# SARS-CoV-2 Total and Subgenomic RNA Viral Load in Hospitalized Patients

**DOI:** 10.1101/2021.02.25.21252493

**Authors:** Derek E. Dimcheff, Andrew L. Valesano, Kalee E. Rumfelt, William J. Fitzsimmons, Christopher Blair, Carmen Mirabelli, Joshua G. Petrie, Emily T. Martin, Chandan Bhambhani, Muneesh Tewari, Adam S. Lauring

## Abstract

Understanding viral load in patients infected with SARS-CoV-2 is critical to epidemiology and infection control. Previous studies have demonstrated that SARS-CoV-2 RNA can be detected for many weeks after symptom onset. The clinical significance of this finding is unclear and, in most patients, likely does not represent active infection. There are, however, patients who shed infectious virus for weeks. Detection of subgenomic RNA transcripts expressed by SARS-CoV-2 has been proposed to represent productive infection and may be a tractable marker for monitoring infectivity. Here, we use RT-PCR to quantify total and subgenomic nucleocapsid (N) and envelope (E) transcripts in 190 SARS-CoV-2 positive samples collected on hospital admission. We relate these findings to duration of symptoms. We find that all transcripts decline at the same rate; however, subgenomic E becomes undetectable before other transcripts. In Kaplan-Meier analysis the median duration of symptoms to a negative test is 14 days for sgE and 25 days for sgN. There is a linear decline in subgenomic RNA compared to total RNA suggesting subgenomic transcript copy number is highly dependent on copy number of total transcripts. The mean difference between total N and subgenomic N is 16-fold (4.0 cycles) and the mean difference between total E and sub-genomic E is 137-fold (7.1 cycles). This relationship is constant over duration of symptoms allowing prediction of subgenomic copy number from total copy number. Although Subgenomic E is undetectable at a time that may more closely reflect the duration of infectivity, its utility in determining active infection may be no more useful than a copy number threshold determined for total transcripts.

## Introduction

The emergence of the severe acute respiratory syndrome virus-2 (SARS-CoV-2) led to the rapid development of diagnostic tests for infection. Many of these tests rely on reverse transcriptase polymerase chain reaction (RT-PCR) to detect total viral RNA. These assays are highly sensitive and specific, with a limit of detection of approximately 5-500 copies of viral RNA per reaction (Vogels et al. 2020). Unfortunately, the relationship between a positive RT-PCR and viral infectivity is unclear, particularly as infection progresses over time. While the median duration of detection of SARS-CoV-2 viral RNA in the upper respiratory tract is roughly 14.5 days from symptom onset, many studies have shown that SARS-CoV-2 RNA can be detected for several weeks, long past when most people are infectious (Walsh et al. 2020).

Because of persistent test positivity, the CDC no longer recommends a test-based strategy for discontinuing isolation precautions. Current guidelines recommend that patients with mild to moderate disease remain isolated for 10 days from symptom onset, while those with either severe disease or an immunocompromising condition remain isolated for 10-20 days from symptom onset. These guidelines are largely supported by studies of viral infectivity in clinical samples over time. (Arons et al. 2020, Wölfel et al. 2020, Bullard et al. 2020) However, recent studies have demonstrated that some immunosuppressed patients may shed infectious virus for weeks, regardless of symptoms (Avanzato et al. 2020, Baang et al. 2020, Choi et al. 2020). Symptom-based strategies for isolation may also be problematic in patients incidentally found to be positive for SARS-CoV-2.

There is no high-throughput, rapid test that distinguishes those who are infectious from those who are not. While viral culture is perhaps the most reliable way to determine infectivity, it is neither timely nor practical in most clinical laboratories. There has been intense interest in whether other molecular markers can be used as correlates of infectivity, including the detection of subgenomic RNA. SARS-CoV-2 has a positive-sense, single-stranded RNA genome of nearly 30 kb. Negative-sense RNA intermediates serve as templates for the synthesis of positive-sense genomic RNA. The viral polymerase also makes subgenomic messages that have a common 5’ leader fused to downsteam open reading frames, which are then translated into nine proteins, including the nucleocapsid (N), spike (S), envelope (E) and membrane (M), and six accessory proteins (3a,6,7a,7b and 8) (Kim et al. 2020). Given this genomic arrangement, subgenomic RNA can be distinguished from total RNA by placement of alternate PCR primers. Because subgenomic transcripts are not packaged into virions, their presence is thought to indicate productively infected cells.

A few studies have evaluated the kinetics of subgenomic RNA during the course of infection. A small study of 9 patients found that no subgenomic RNA could be found in the throat up to 5 days post symptom onset. (Wölfel et al. 2020) A larger study of 35 patients showed that subgenomic RNA and culturable virus were not detectable after 8 days from symptom onset in mild COVD-19 disease, but total SARS-CoV-2 transcripts persisted for weeks (Perera et al. 2020). Less is known about subgenomic RNA kinetics in hospitalized patients with severe disease and in immunosuppressed patients.

To determine the utility of subgenomic RNA as a correlate of SARS-CoV-2 infectivity, we used RT-PCR to amplify the total and subgenomic nucleoprotein gene (N) and envelope gene (E) from 190 individual patient samples collected on admission to the hospital. We show that subgenomic transcripts become undetectable before total transcripts when evaluated in relation to duration of symptoms. Because expression levels of total and subgenomic transcripts are highly correlated, the added benefit of measuring subgenomic RNA is limited.

## Materials and Methods

### Samples and participants

Nasopharyngeal (NP) samples were obtained from patients admitted to the University of Michigan between March 13, 2020 to June 10, 2020. Dacron swabs were placed in 3 ml viral transport media and transported to the clinical microbiology laboratory for testing. Residual samples were stored in a central biorepository at −80°C. In total, 185 patients with RT-PCR confirmed SARS-CoV-2 and clinical information were included in our study. An additional five patients without clinical data were included for a total of 190 individual NP swabs. Clinical data, including date of symptom onset, basic demographic data, and outcome were abstracted from clinician notes. This study protocol was reviewed and approved by the University of Michigan Institutional Review Board (IRB). The IRB determined the study to be exempt from requirement for informed consent given retrospective nature of this study and this use of stored biospecimens and deidentified data.

### Quantitative RT-PCR

Residual samples from NP swabs were centrifuged at 1200 x g and 200 µl of sample used for RNA extraction. RNA was extracted with the Invitrogen PureLink Pro 96 Viral RNA/DNA Purification Kit (Invitrogen, Carlsbad, CA). Samples were eluted in 100 µl and stored at −80C.

Amplification of total and subgenomic transcripts for nucleocapsid (N) and envelope (E) genes was performed using conditions outlined in the CDC 2019-Novel Coronavirus EUA protocol (https://www.fda.gov/media/134922/download). We diluted our clinical samples 1:5 in nuclease free H_2_O and used 5 µl of this dilution for 1 µl equivalent total RNA per reaction. This was done to preserve clinical sample, and the dilution was accounted for in copy number calculations. Reactions were preformed using Taqpath 1-step RT-qPCR master mix (Thermofisher, Waltham, MA), 500 nM of each primer, and 250 nM of each probe in a total reaction volume of 20 µl. Cycling conditions were as follows: activation of uracil-N-glycosylase for 2 min at 25° C, reverse transcription for 15 min at 50° C, denaturation for 2 min at 95° C, and 45 cycles of 3 s at 95° C, 30 s at 55° C on an Applied Biosystems 7500 FAST real-time PCR system. Cycle threshold (Ct) was determined uniformly across PCR runs.

Subgenomic transcripts were amplified by substituting subgenomic leader sequence sgLeadSARSCoV2-F: 5’-CGATCTCTTGTAGATCTGTTCTC-3’ (Wölfel et al. 2020) as the forward primer for E or N together with the reverse primers and probes for N and E genes. For total E, the primers and probes were from the Charite/Berlin protocol (Corman et al. 2020) and the sequences are as follows: E_Sarbeco_F: ACAGGTACGTTAATAGTTAATAGCGT; E_Sarbeco_R: 5-ATATTGCAGCAGTACGCACACA-3; E_Sarbeco_P1: FAM-ACACTAGCCATCCTTACTGCGCTTCG-3IABkFQ. The N gene was amplified using the CDC N1 primer and probe set as follows: 2019-nCoV_N1 Forward Primer GACCCCAAA ATCAGCGAAAT; 2019-nCoV_N1 Reverse Primer TCTGGTTACTGCCAGTTGAATCTG; 2019- nCoV_N1 Probe ACCCCGCATTACGTTTGGTGG ACC. Probe sequences were FAM labeled with Iowa Black quencher (Integrated DNA Technologies, Coralville, IA).

### Droplet Digital PCR

Absolute copy numbers of N, E, sgN and sgE were determined by droplet digital PCR (ddPCR) using SARS-CoV-2 RNA as template from HuH-7 infected cells. HuH-7 cells were infected with SARS-CoV-2 strain WA-1. RNA was harvested at 48 hours post infection using trizol and Zymo columns. ddPCR was performed using the QX200 Droplet Digital PCR System (Bio-Rad, Hercules, CA). Sample reaction contained HuH-7 RNA/sgRNA template (5.5 ul), the respective forward and reverse primers (final concentration 900nM each) and the respective FAM labeled probe (final concentration 250nM). The other reaction components common to all reactions were added at the final concentration as per the manufacturer’s recommendation (Bio-Rad # 1864021; 1X Supermix, 20 units/ul Reverse Transcriptase and 15 mM DTT). Each 20 ul reaction mix was partitioned into droplets using the QX200 droplet generator (Bio-Rad), transferred into a 96-well plate, sealed and cycled in a C1000 Thermal Cycler (Bio-Rad). The cycling conditions used were as per the manufacturer’s recommendation (Bio-Rad’s expert design assay for SARS-CoV-2 # dEXD28563542; hold at 25°C for 3 min, reverse transcription at 50°C for 60 min, enzyme activation for 10 minutes at 95°C for 1 cycle, denaturation at 94°C for 30 seconds and annealing/extension at 60°C for 60 seconds for 40 cycles, enzyme deactivation for 10 minutes at 98°C for 1 cycle followed by a hold at 4°C). All steps were performed with a 2°C/second ramp rate and the lid temperature set at 105°C. Droplets were read using QuantaSoft Software in the QX200 reader (Bio-Rad). Once copy number was determined, a ten-fold dilution series (from 1×107 copies to 10 copies per 20ul reaction) of this sample was performed to generate standard curves and used to determine copy number of clinical samples. Copy number was corrected for dilution of sample to reflect copies per milliliter of viral transport media from original NP swab sample.

### Statistical Analyses

Continuous variables were expressed as medians and interquartile ranges (IQRs) or means and standard deviations. Simple linear regression and Kaplan-Meier analysis were performed using SPSS software version 27 (IBM Corporation) and GraphPad (Prism). Statistical significance was set as P <0.05.

## RESULTS

### Patient characteristics

We obtained NP swabs from 190 SARS-CoV-2 positive patients admitted to the University of Michigan Hospital from March 13, 2020 to June 10, 2020. We were able to obtain clinical information from 185 of the 190 patients. In our sample, 56.8% were men (105/185). The median age was 64 (IQR: 53-75) years old. The median days from symptom onset to SARS-CoV-2 testing was 5 days (IQR 3-8). Three patients reported no symptoms consistent with COVID-19 infection on admission but reported symptoms later in their hospital course.

### Symptom duration compared to total and subgenomic transcripts

We found that there was no difference in the rate of decline of SARS-CoV-2 transcripts when compared to symptom duration with all slopes equal (p>0.05) (Figure 1). We did however find a significant difference in the X-intercepts for the regression line of viral RNA transcript decline over time. There was no difference in intercepts between total E and N (p>0.72). When total N was compared to sgN the X-intercepts were significantly different (P<0.0001) as was the comparison of E to sgE (P<0.0001). Finally, there was a significant difference between intercepts for subgenomic transcripts sgN and sgE (P<0.001). The differences X-intercepts reflects relative differences in days until RNA is no longer detectable based on a threshold of 40 cycles.

**Figure 1.**
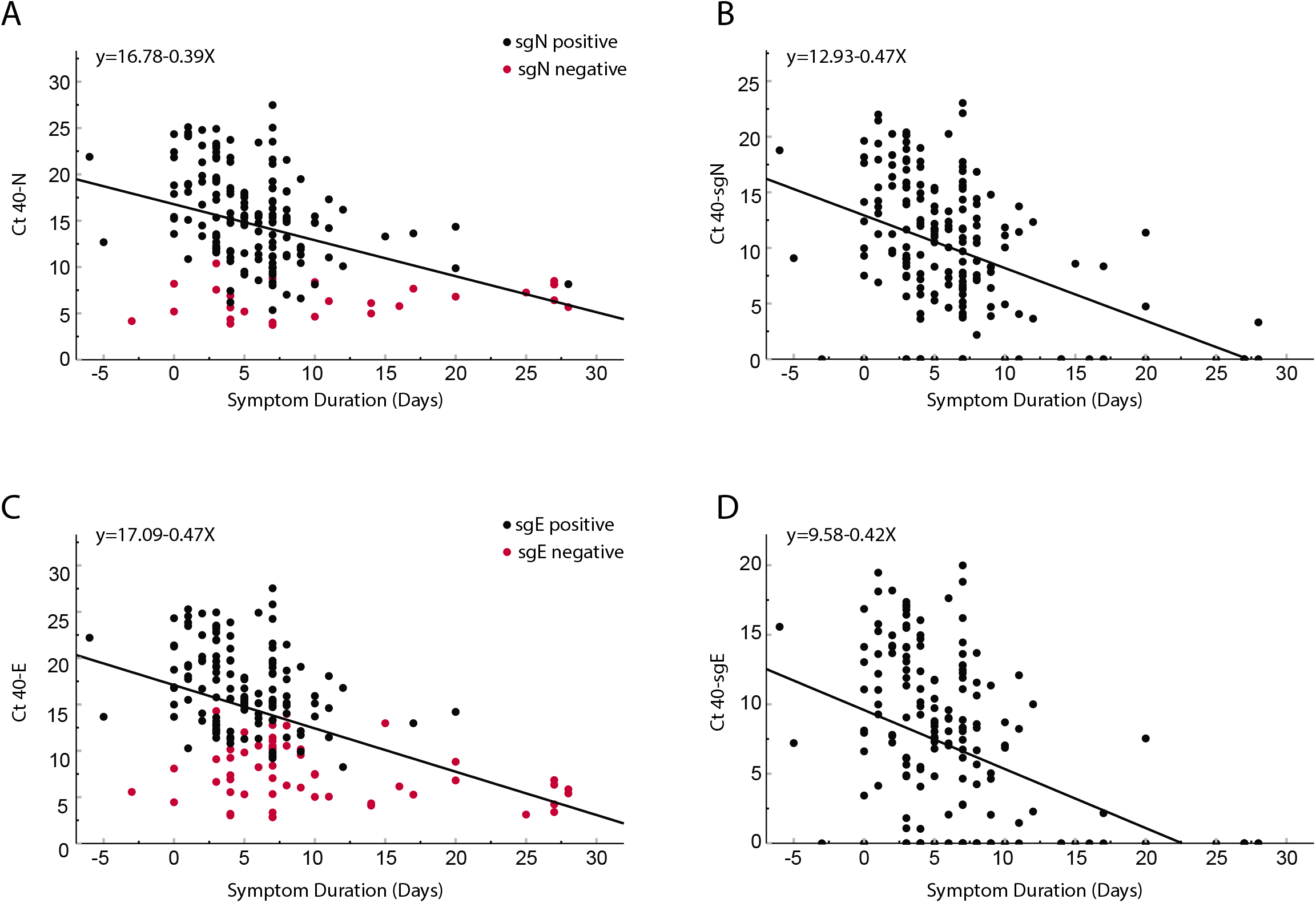
Comparison of cycle threshold versus day from symptom onset for clinical samples obtained from 185 inpatients. Total N (Panel A), subgenomic N (Panel B), total E (Panel C) and subgenomic E (Panel D). Red dots in panel A and C represent subgenomic negative samples and black dots represent subgenomic positive samples. Of the 185 patients, 56 were negative for sgE and 28 were negative for sgN (Shown on y-axis). Pearson correlation coefficients: N = −0.404 p<0.0001; SgN = −0.466, p<0.0001; E = −0.456 p<0.0001; sgE = −0.427 <0.0001. Linear regression line equations are indicated in each panel.

We used a Kaplan-Meier analysis to further investigate the relationship between symptom duration and a negative subgenomic RNA test. The median duration from symptom onset to a negative sgN RT-PCR was 25 days (95% CI: 19.9-30.1) and 14 days for sgE (95% CI 9.6-18.4 days). This difference was statistically significant (P=0.001) (Figure 2). All 185 patients were positive for total RNA regardless of symptom duration. By ≥15 days from symptom onset, only 14% of patients were positive for sgE compared to 35.7% for sgN.

**Figure 2.**
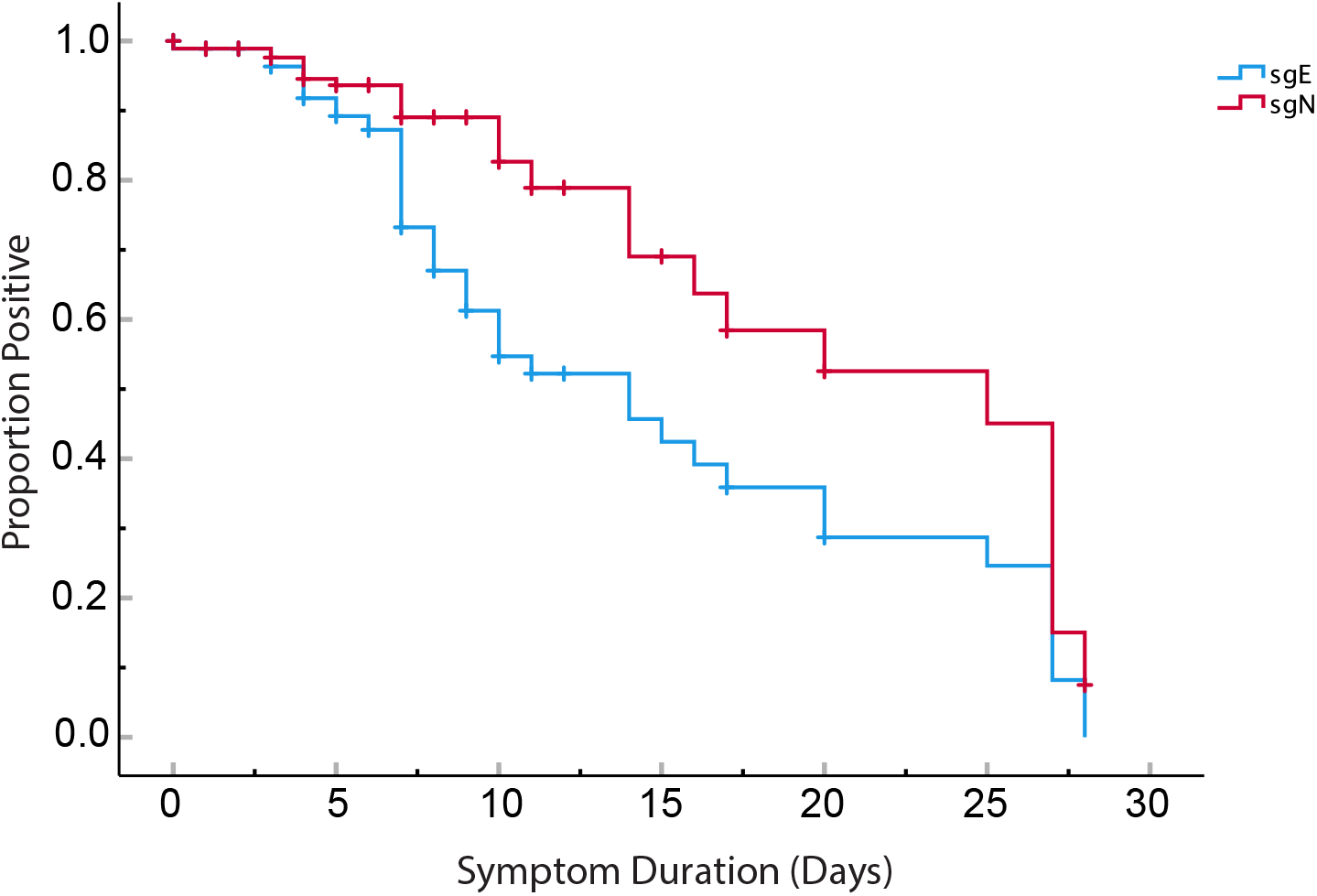
Kaplan-Meier analysis showing proportion of patients with positive RT-PCR for subgenomic transcripts versus day from symptom onset. The median duration from symptom onset to a negative sgN RT-PCR was 25 days (95% CI: 19.9-30.1) and 14 days for sgE (95% CI 9.6-18.4). This difference between the curves was significant P=0.001. Vertical hash marks indicate censored cases.

Recent studies have demonstrated that the total copy number threshold that correlates with a loss of culture infectivity is between 5.5 and 6.5 log_10_ copies per ml (Wölfel et al. 2020, Bullard et al. 2020, van Kampen et al. 2021). Primers used in this study amplify total RNA (genomic plus subgenomic RNA), and by pairing the subgenomic leader primer with reverse E and N primers distinguish subgenomic from total RNA. We determined copy number in clinical samples using standard curves generated with known copy number from ddPCR experiments for total N, total E, sgE and sgN. At 6.5 log_10_ total E copies, 47% (56/119) of samples were negative for sgE. At a total N copy number of 6.5 log_10_, 24% (28/119) were negative for sgN. There we no samples negative for subgenomic at a copy number greater than 6.5 log_10_ total copies per ml (Supplementary Figure 1).

### Relationship of total to subgenomic transcripts

To further understand the relationship between total and subgenomic transcripts for the N and E genes, we compared Ct values from 190 clinical samples. We found that 162 of 190 (85%) patients had both a detectable N and sgN, and 132 (69%) patients had both a detectable E and sgE. Subgenomic transcripts declined linearly along with total RNA transcripts (Supplementary Figure 2). There was no difference in the slope of subgenomic to total RNA regression (linear regression, slope of N vs sgN −0.99 and slope of E vs sgE −1.03 P=0.09). We were unable to detect sgE RNA transcripts once total E reached 32 cycles, and were unable to detect sgN transcripts once total N reached 35 cycles.

We found there was very little difference between expression levels of total N and E. The mean Ct value of E was 25.76 (STD 5.69) and N was 25.6 (STD 5.6 cycles). In cell culture and in clinical samples, we found that total RNA levels were higher than subgenomic RNA levels. Total N was expressed at a 4.5-fold (2.2, STD 0.24 cycles) higher level than sgN, and E was expressed at a 13.9-fold (3.8, STD 0.2 cycles) higher level compared to sgE in HuH-7 cells at 48 hours post infection (Figure 3). This difference was more dramatic in clinical samples. Total N was expressed at a 16-fold (4.0, STD 1.1 cycles) higher level than sgN transcripts and E was expressed at a 137.2-fold (7.1, STD 1.3 cycles) higher level than sgE when all clinical samples were compared together (Figure 3). This relationship remained unchanged throughout the course of infection (Figure 3). Furthermore, the large fold difference in subgenomic compared to total transcripts indicates that subgenomic RNA contributes little to the overall signal of total transcripts.

**Figure 3.**
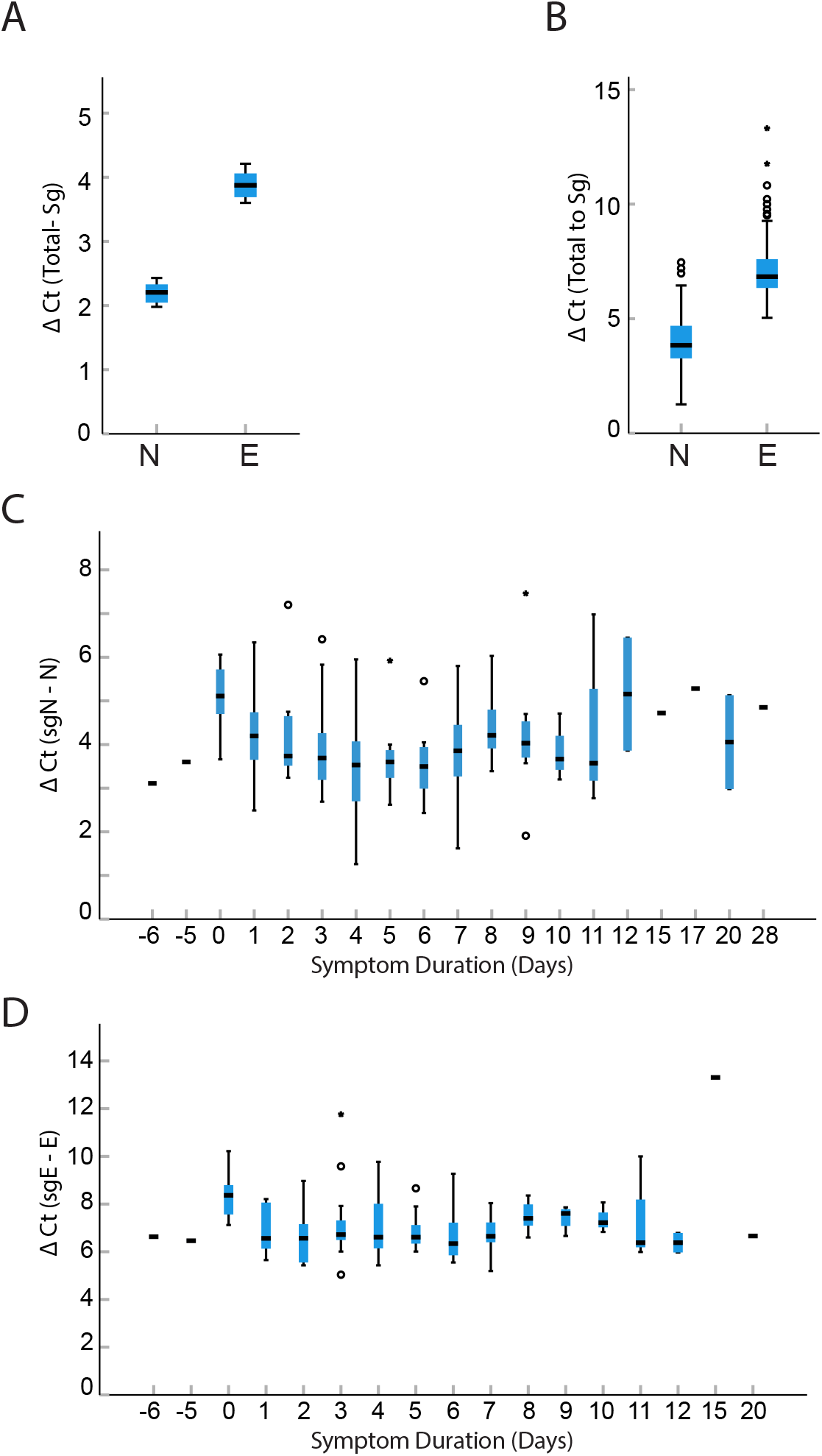
Box plots comparing difference in cycle threshold values, delta Ct (subgenomic Ct-genomic Ct) for cell culture and clinical samples. Total N is expressed at a 4.5-fold (2.2 cycles, STD 0.24) higher level than sgN and total E is expressed at a 13.9-fold (3.8 cycles STD 0.2) higher level compared to sgE in HuH-7 cells (Panel A). The difference was more dramatic in NP samples from inpatients where N was expressed at 16-fold (4.0 cycles, STD 1.1) greater than sgN and E was expressed at 137.2-fold (7.1 cycles, STD 1.3) greater than sgE expression (Panel B). This relationship was found at all times post symptom onset (n=185) for N (Panel C) and E (Panel D). Horizontal hash represents median value, if no IQM shown then N=1 for that sample. Moderate and extreme outliers noted by circles and stars respectively.

### Subgenomic RNA in a persistently infected patient

Emerging data indicates that some immunosuppressed patients can be persistently positive for SARS-CoV-2 by RT-PCR and shed infectious virus for weeks. We evaluated total and subgenomic RNA from a patient persistently infected with SARS-CoV-2 for 119 days. This patient showed continued expression of total and subgenomic transcripts throughout this time. The mean fold difference of total N to subgenomic N was 11.3-fold (3.5 cycles) and the mean fold difference of total E to subgenomic E was 84-fold (6.4 cycles) (Figure 4). This ratio was maintained over the 119 day course.

**Figure 4.**
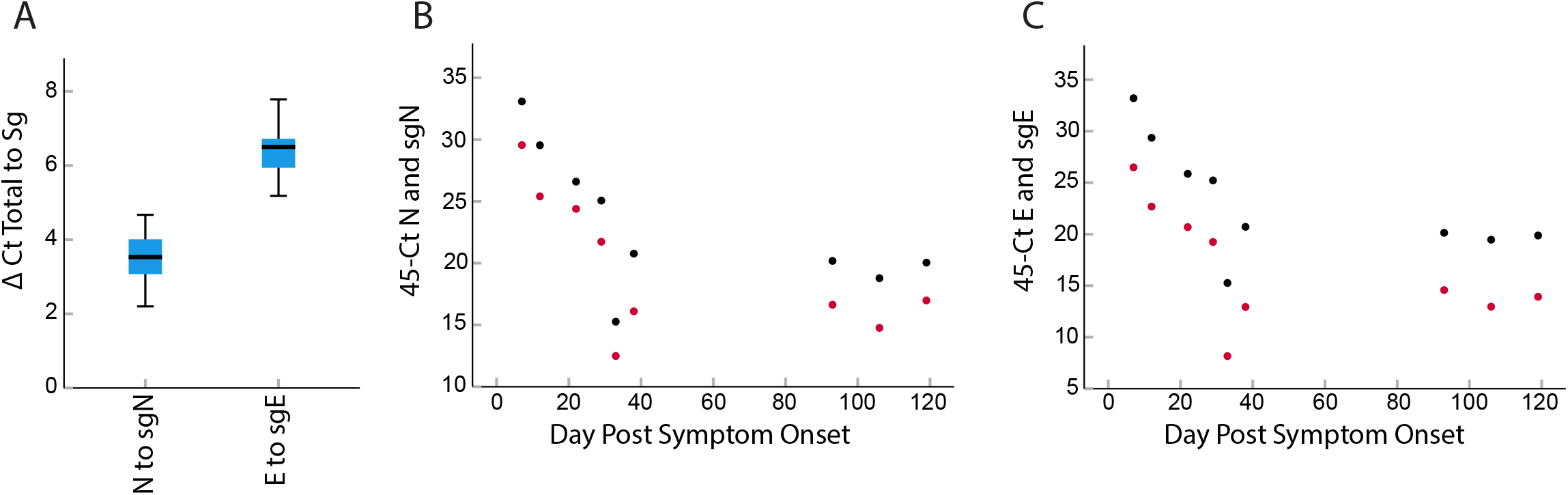
Relationship between total and subgenomic RNA in a persistently infectious patient. Total N was expressed at a 11-fold (3.5 cycles STD 0.75) higher level than subgenomic N, and total E was expressed at a 84-fold (6.4 cycles, STD 0.8) higher level than subgenomic E when all time points were combined (Panel A). Time course showing 45 minus cycle thresholds for total (black) and subgenomic RNA (red) over time. This patient showed persistently positive total and subgenomic RNA for 119 days. The ratio of total N and sgN of 3 to 4 cycles was maintained over the duration of infection (Panel B). This was also observed for E and sgE with the ratio of 6 to 7 cycles maintained thorough the course of infection (Panel C).

## DISCUSSION

In this study, we found that subgenomic transcripts become undetectable at an earlier timepoint after infection than total transcripts. The median duration of symptoms until a negative RT-PCR test was 14 days for subgenomic E and 25 days for subgenomic N. Total RNA was positive at all time points in our study up to 28 days. We found a strong correlation between total and subgenomic transcript levels. They decline at the same rate and have a fixed ratio at all time points post symptom onset. The predictability of subgenomic copy number from total copy number indicates that detection of subgenomic RNA does not add additional information about infectivity that cannot be gained from total copy number alone.

A recent study in hospitalized patients showed that fewer than 5% of patients are infectious 15 days after the onset of symptoms (van Kampen et al. 2021). We find that subgenomic E becomes undetectable at a median of 14 days and only 14% (2/14) of patients were positive for subgenomic E after this time point. Our results suggest that the disappearance of subgenomic E RNA may correlate with a time when patients are no longer infectious.

Despite our finding that a negative subgenomic E RT-PCR seems to correlate with a time when patients are not infectious, the supposition that any negative subgenomic RNA transcript can be used to predict infectivity is problematic. In our analysis, the median number of days from symptom onset to a negative subgenomic N RT-PCR was 25 days. Using subgenomic N as a marker for infectivity would significantly prolong isolation and not predict infectivity. This difference is likely due to the higher levels of expression of subgenomic N relative to subgenomic E.

A negative subgenomic RNA test likely indicates that patients are not infectious simply because a negative subgenomic RNA test correlates with a low total RNA copy number. Previous studies have shown a copy number below 6.5 log_10_ copies per ml is a threshold for culturable virus. In fact, we found that all negative subgenomic transcripts were in samples with total copy number below this threshold. However, we also find many samples that are below 6.5 log_10_ copies per ml, but still express subgenomic E and N. This is in line with a recent study that showed subgenomic RNA could be detected from samples where infectivity in cell culture was negative (van Kampen et al. 2021). It is likely that subgenomic RNA also has a copy number threshold below which infectious virus cannot be isolated. Because of the strong correlation in copy number of total and subgenomic transcripts throughout the course of infection and in a persistently infected patient, we do not believe that subgenomic RNA provides additional information not gained by total copy number.

Previous research suggested that subgenomic RNA is a suitable marker for active infection because it degrades more rapidly than total RNA. One study inoculated animals with infectious and inactivated SARS-CoV-2, which had high levels of total and subgenomic RNA. Total RNA copy number on day 1 post infection from inactivated virus ranged from roughly 4 log_10_ to 6 log_10_ copies per ml but no subgenomic E RNA was detected. This was interpreted to mean that subgenomic RNA rapidly degrades without replication, but total RNA does not. (Speranza et al. 2020) In our study, at a total copy number of 4 log_10_ no subgenomic E could be detected, and at less than 6 log_10_ per ml of total RNA 47% (56/119) of samples were negative for subgenomic E. We suspect this finding is not due to differential degradation, but rather to differential detection of transcripts. Furthermore, if there was a difference in rates of degradation, we would expect to see the ratio of total RNA to subgenomic RNA increase overtime, which we did not see.

Although there is emerging data supporting isolation guidelines in mild/moderate and severe COVID-19 infection, little is known about infectivity in immunosuppressed patients. In this study we showed persistent total and subgenomic RNA expression up to 119 days with a previous study demonstrating infectious virus could be isolated through this course (Baang et al. 2020). Because low levels of subgenomic RNA can be detected in the absence of infectiousness, we do not endorse using subgenomic RNA to determine infectivity in these patients. As with non-immunosuppressed patients, a copy number threshold of total RNA would likely provide the same information which can be gained from copy number on total RNA.

Our study is limited by the fact that we did not attempt to isolate virus in culture from clinical samples and correlate to total or subgenomic copy number. We did however investigate correlation of transcripts to symptom duration, which has been used as a benchmark for infectivity. We are aware that recall bias may affect reported symptom duration particularly in ill hospitalized patients. This is mitigated to some degree by our large sample size. Differences in copy number observed could be the result of differences in efficiency of RT-PCR reactions across total and subgenomic transcripts. We confirmed differences in copy number using absolute copy number determined using standard curves generated with a known copy number using ddPCR for N, E, sgE and sgN. Furthermore our findings are in agreement with direct RNA sequencing studies showing the subgenomic N is expressed at higher levels than subgenomic E. (Kim et al. 2020)

In this study of inpatients infected with SARS-CoV-2, we find that subgenomic E transcripts become undetectable before other transcripts largely because it is transcribed at far lower levels than other transcripts studied here. There is a fixed relationship between copy number of total and subgenomic transcripts over the course of infection. Because of this consistent relationship between total and subgenomic RNA, we do not believe subgenomic RNA detection adds additional information regarding infectivity that cannot be gained from copy number threshold validated to infectivity determined by cell culture.

## Data Availability

All available data are presented in the manuscript.

## ACKNOWLEDGEMEMTS

We thank the University of Michigan Clinical Microbiology Laboratory and the University of Michigan Central Biorepository for their assistance in providing samples. This work was supported by a University of Michigan COVID-19 Response Innovation Grant (to ASL), K01AI141579 (to JGP) and CDC U01 IP000974 (to ETM)

## FIGURE LEGENDS

**Supplementary Figure 1.**
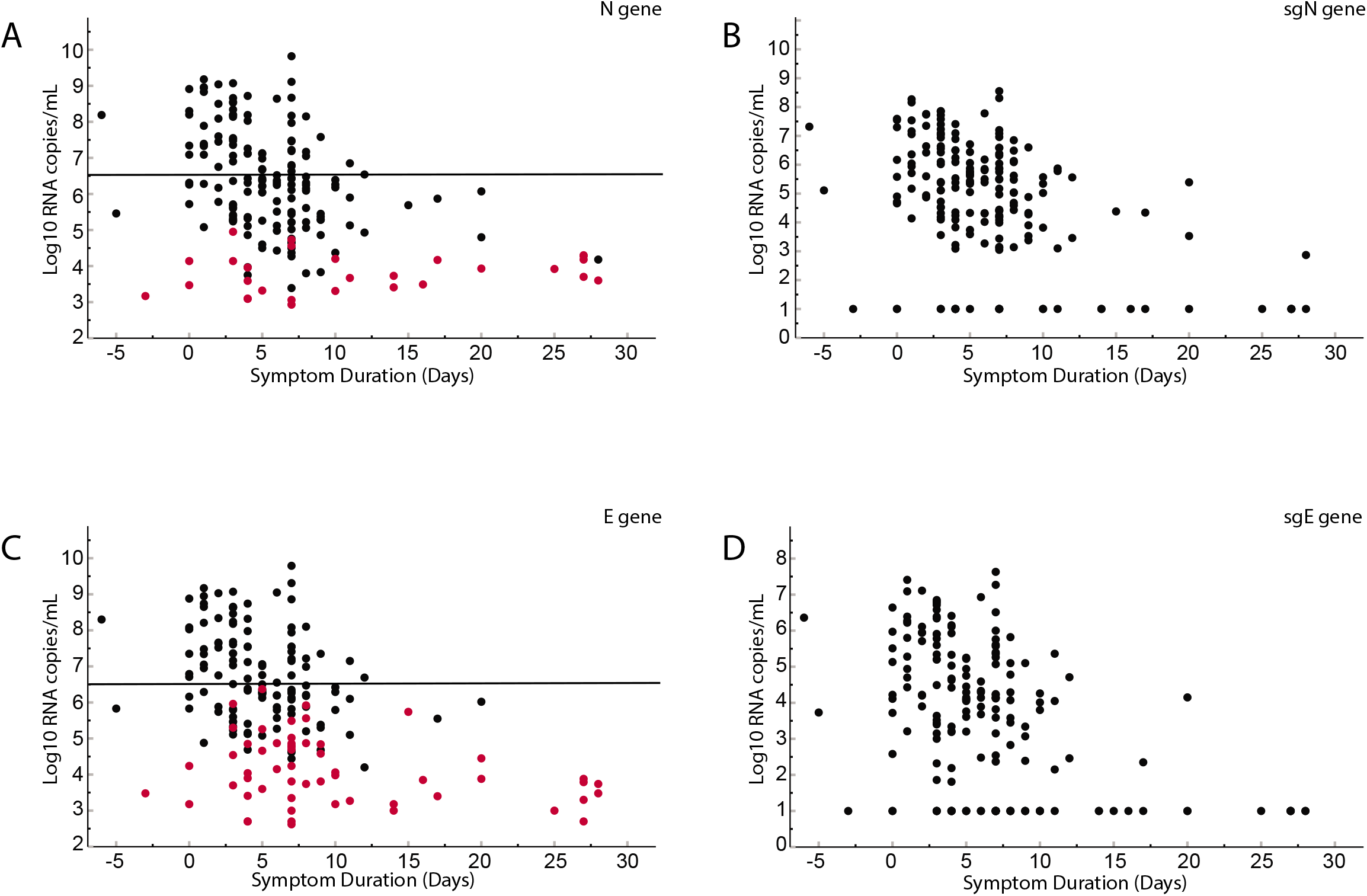
Comparison of viral load in log_10_ copy number per ml from NP swabs of 185 patients versus duration of symptoms. Panel A shows total N, Panel B shows subgenomic N, Panel C shows total E, Panel D shows subgenomic E. Line at 6.5 log_10_ is the threshold for infectivity found in previous studies. In Panels B and D, samples that were undetectable after 45 cycles were arbitrarily set to 1 log_10_ per ml. Red circles in panels A and B represent the total copy number of samples that were negative for subgenomic RNA. Subgenomic E is negative at a higher total copy number than subgenomic N.

**Supplementary Figure 2.**
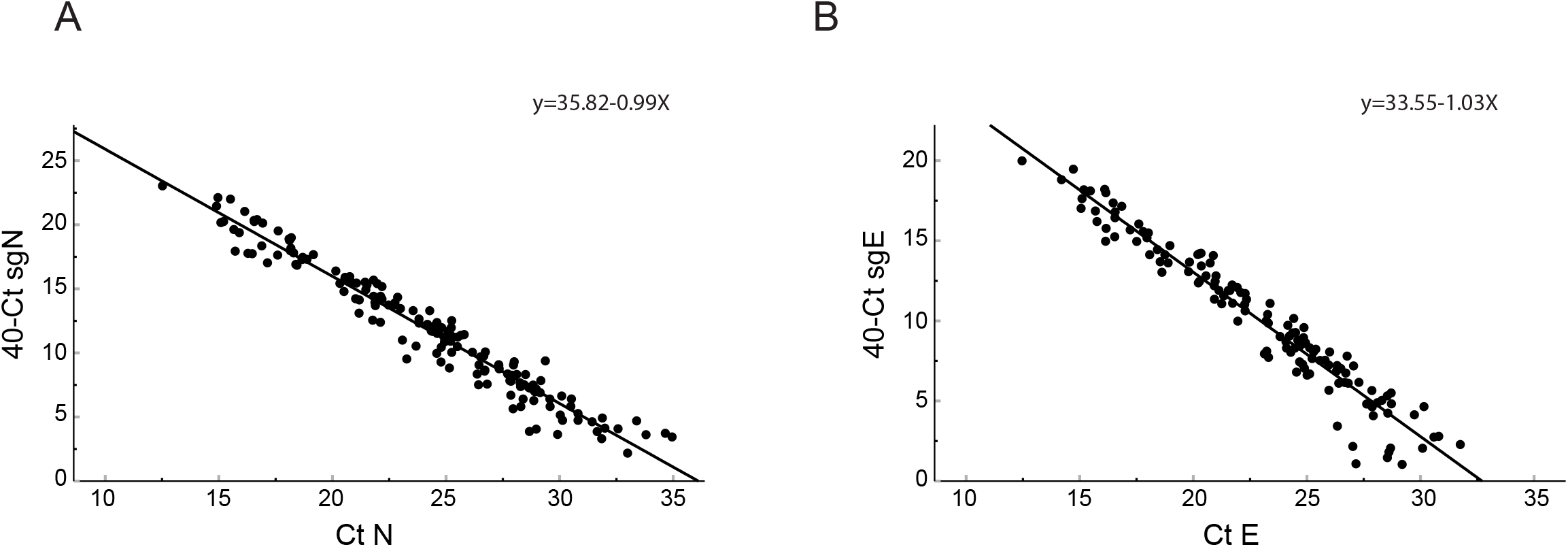
Comparison of cycle thresholds for subgenomic RNA to total RNA for subgenomic N and total N (Panel A) and subgenomic E and total E (Panel B). Samples were obtained from patients admitted to the hospital (n=190). Subgenomic Ct values on the y-axis are represented as 40 cycles minus the Ct value of subgenomic RNA meaning a lower Ct value represents a lower copy number of subgenomic RNA. Increasing Ct values on the x-axis for total RNA represent decreasing copy number for total RNA. Linear regression analysis showed no difference in the slope between the two lines. There was a significant difference between X-intercepts. The X-intercept for the comparison of Ct value of N vs Ct value of sgN was 36.01 (CI 35.67 to 36.57) and the X-intercept for the comparison of Ct value of E vs Ct value of sgE was 32.71 (CI: 32.25 to 33.2) (P<0.0001). Pearson correlation coefficient for sgN/N = −0.976, P<0.0001; for sgE/E = −0.969 (p<0.0001).

